# The Use of Carotid Web Angioarchitecture in Stratification of Stroke Risk

**DOI:** 10.1101/2024.11.07.24316945

**Authors:** Bruck Negash, Daniel D. Wiggan, Eric A. Grin, Karl L. Sangwon, Charlotte Chung, Eleanor Gutstadt, Vera Sharashidze, Eytan Raz, Maksim Shapiro, Koto Ishida, Jose L Torres, Cen Zhang, Michelle A Nakatsuka, Sara K Rostanski, Melissa J. Rethana, Alexandra Kvernland, Matthew Sanger, Kaitlyn Lillemoe, Alexander Allen, Sean Kelly, Jacob Baranoski, Caleb Rutledge, Howard A. Riina, Peter Kim Nelson, Erez Nossek

## Abstract

**Objective:** To validate the carotid web (CW) risk stratification assessment described in our previous work with a larger cohort of patients with incidentally found asymptomatic CWs.

**Methods:** A retrospective analysis of our electronic medical record database identified all patients with a diagnosis of CW from 2017-2024 at our institution. We included symptomatic patients and those with asymptomatic carotid webs, meaning patients with incidentally found webs and no history of stroke or transient ischemic attack (TIA). Patient charts were reviewed for demographics, imaging, comorbidities, hospital course, and incidence of stroke after diagnosis of an asymptomatic carotid web. All angles were measured as described in our previous work on a sagittal reconstruction of CTA imaging of the neck in which the common carotid artery (CCA), external carotid artery (ECA), and ICA could be well visualized, along with the CW itself. A standard logistic regression was performed to evaluate the association between the number of high-risk angles and stroke risk.

**Results:** 26 asymptomatic and nine symptomatic patients were identified. Patients were categorized as having 0, 1, or 2+ high-risk angle values. Patients with more high-risk angles had a markedly higher risk of stroke (OR = 5.91, 95% CI: [4.25, 8.24]). The probability of stroke increased with the number of high-risk angles: patients with 2+ high-risk angles (83.4%) had the highest stroke probability compared to those with 0 (2.8%) or 1 (27.7%) high-risk angles. In the asymptomatic cohort, mean angles all fell below the high-risk threshold values. In the symptomatic cohort, mean angles were above the high-risk threshold values, whereas the mean CPT (53.6°) angle fell below the cut-off value for designation as a high-risk angle.

**Conclusions:** Given the successful stratification of CWs into high and low-risk groups in this study, the utilization of geometric CW parameters may play a crucial role in improvement of patient selection for intervention in patients with an incidental diagnosis of CW.

## Introduction

Carotid webs (CWs) are endoluminal bands of fibrous tissue extending from the posterior margin of the internal carotid artery (ICA) just distal to the carotid bulb.^1^ Notably, CWs have been implicated as a cause of thromboembolic stroke, particularly in younger patients lacking other identifiable stroke risk factors.^2,3^ While CWs themselves are rare, with a reported prevalence of 2.3%, as awareness of CWs rises, increasing numbers of patients are being incidentally diagnosed.^4^ This number will likely continue to rise considering the steadily increasing rate of medical imaging in healthcare.^5^

CWs can be categorized as symptomatic, causing a stroke or transient ischemic attack (TIA), or asymptomatic, presenting as an incidental finding.^6^ To date, no prior research has determined whether a pathophysiological or structural difference exists between symptomatic and asymptomatic webs. In this manner, physicians currently have no way to assess whether a patient with an incidentally discovered CW is at risk of having a future stroke. Given the relatively high incidence of incidentally discovered asymptomatic ACWs, determining future stroke in patients with asymptomatic CWs would be of great aid to clinicians in determining the optimal course of management.

Our previous work proposed a new method by which to stratify stroke risk in CW patients according to anatomic data on the ICA, common carotid artery (CCA), and CW (Table 1).^7^ This is accomplished by considering the web’s key structural features and relationship to the carotid bifurcation, a collection of findings we name the carotid web angioarchitecture.

**Table 1.** Angioarchitectural parameters and risk thresholds.

| Angle | Angle Description | Risk Threshold |
| --- | --- | --- |
| IWP | Angle between the Superior Pouch-Wall line and the Longitudinal Internal Carotid Artery line | $\geq 92.4^{\circ}$ |
| CWP | Angle between the Superior Pouch-Wall line and the Longitudinal Common Carotid Artery line | $\geq 41.7^{\circ}$ |
| IPT | Angle between the Pouch-Bifurcation line and the Longitudinal Internal Carotid Artery line | $\geq 85.7^{\circ}$ |
| CPT | Angle between the Pouch-Bifurcation line and the Longitudinal Common Carotid Artery line | $\geq 89.4^{\circ}$ |

In this current study, we aimed to validate this CW angioarchitecture-based risk stratification assessment within a new population of patients including a larger cohort with incidentally found, asymptomatic CWs.

## Methods

A single-institution study was performed. Patient consent was waived per the institutional review board (IRB s22-00111). Our electronic medical record database was queried for all imaging impressions containing the word “carotid web” over the seven-year period from 2017-2024. Patients were identified to have CWs based on classic findings on CTA, including a radiographic “shelf-like” filling defect at the posterior wall of the proximal internal carotid artery (ICA) and description by the diagnosing neuroradiologist.

Patient files were then further reviewed for ischemic signs and symptoms. Symptomatic patients were defined as patients with a diagnosis of CW and a history of ipsilateral stroke or TIA. Diagnoses of stroke or TIA were based on neurologic symptoms, neuroimaging using CT or MRI, and vessel imaging using CTA or angiography. All diagnoses of stroke or TIA were made by a board-certified vascular neurologist. Further, all strokes occurred ipsilateral to the identified carotid web, and in each case, the carotid web was determined by the vascular neurology team to be the most likely cause of stroke after a thorough assessment ruling out other causes of ischemic events. Asymptomatic patients were defined as patients with CWs on CTA imaging who had no history of stroke or TIA. These patients were incidentally found to have CWs while undergoing imaging for unrelated conditions or circumstances.

Patient charts were then reviewed for demographics, imaging findings, clinical course, and outcomes after CW diagnosis. Angioarchitectural parameters, including the ICA – web pouch angle (IWP), CCA -- web pouch angle (CWP), ICA -- pouch tip angle (IPT), and CCA -- pouch tip angle (CPT), were measured as described in our previous work and serve as proxies for flow dynamics around the CW (Figure 1).^7^ These features were measured consistently by the same two investigators. All angles were measured on a sagittal reconstruction of CTA imaging of the neck in which the CCA, external carotid artery (ECA), and ICA could be well visualized, along with the CW itself.

**Figure 1.**
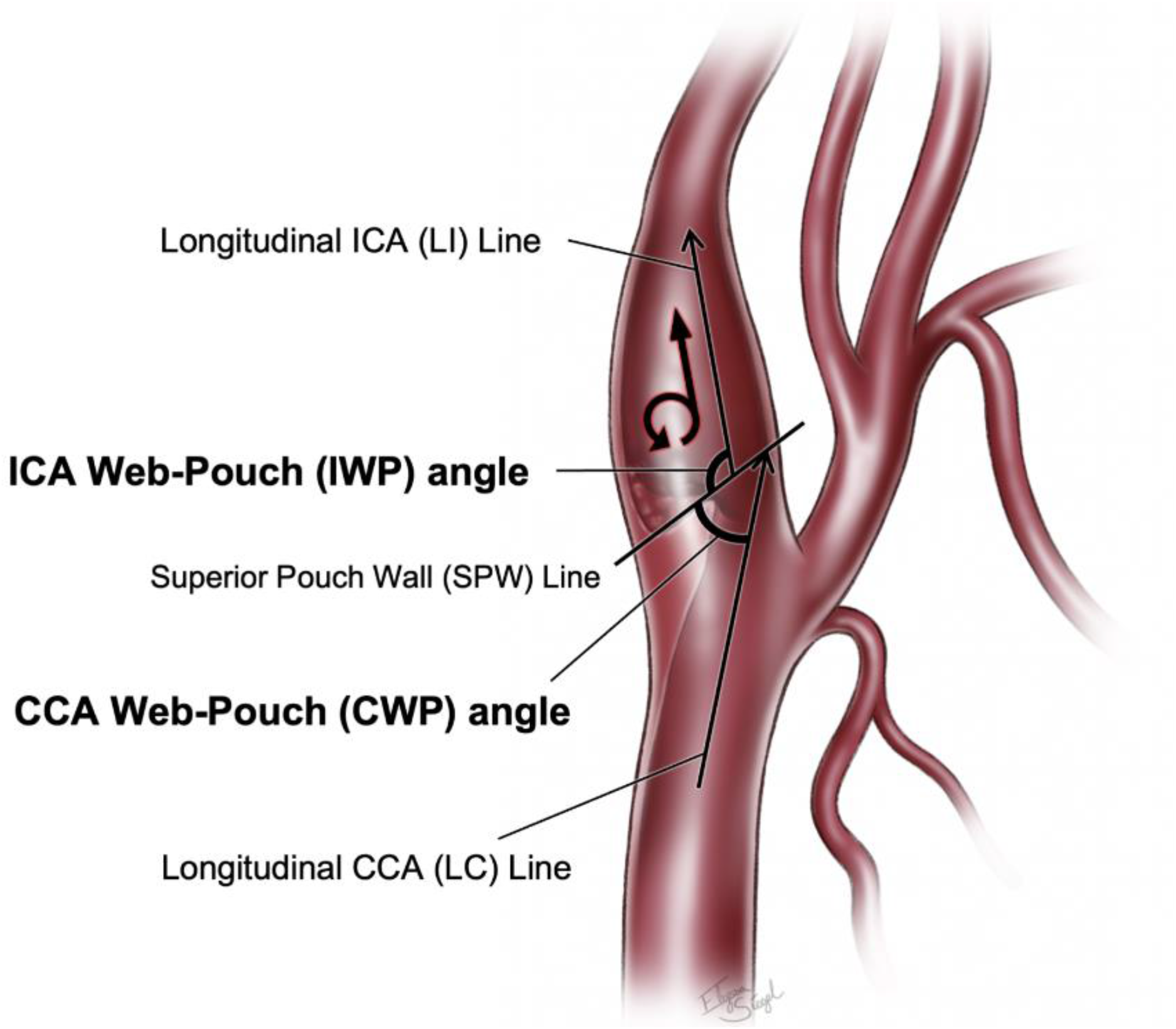
Illustrations demonstrating the (A) ICA Web-Pouch (IWP) angle, CCA Web-Pouch (CWP) angle, (B) ICA Pouch-Tip (IPT) angle, and CCA Pouch-Tip (CPT) angle.

For statistical analysis, each patient, and thus each CW, was then categorized as having either 0, 1, or 2+ high-risk angle values, meaning values above the predetermined thresholds for each parameter. We performed a standard logistic regression to evaluate the association between the number of high-risk angles and stroke risk. The number of high-risk angles was grouped into three categories: 0, 1, and 2+. The outcome was binary (stroke occurrence), where 1 represented stroke and 0 represented no stroke. The predictor variable was standardized prior to modeling to improve convergence. Odds ratios (OR) and 95% confidence intervals (CI) were calculated to quantify the relationship between high-risk angles and stroke risk. The predicted probability of stroke was plotted for each group. STROBE reporting guidelines were adhered to throughout our study.^8^

## Results

Twenty six asymptomatic and nine symptomatic patients with a mean age at diagnosis of 57.4 years (range 28 -80 years) and 60.8 years (range 32 - 89 years) respectively. No significant differences in age, sex, history of hypertension, history of hyperlipidemia or smoking status were observed between the two cohorts (Table 2).

**Table 2.** Demographics of patient population.

|  | Symptomatic n (%) | Asymptomatic n (%) |
| --- | --- | --- |
| Comorbidities |  |  |
| Hypertension | 9(75) | 14(51) |
| Hyperlipidemia | 11(92) | 12(44) |
| Smoking History | 5(42) | 13(48) |
| Race |  |  |
| Black | 2(17) | 4(15) |
| White | 6(50) | 15(56) |
| Asian | 3(25) | 2(7) |
| Hispanic | 1(8) | 1(4) |
| Unknown | 0(0) | 5(18) |
| Sex |  |  |
| Male | 4(33) | 12(44) |
| Female | 8(66) | 15(56) |

Logistic regression revealed a significant positive association between the number of high-risk angles and stroke risk (Figure 2). Patients with more high-risk angles had a markedly higher risk of stroke, with each additional high-risk angle associated with a significant increase in the odds of stroke (OR = 5.91, 95% CI: [4.25, 8.24]).

**Figure 2.**
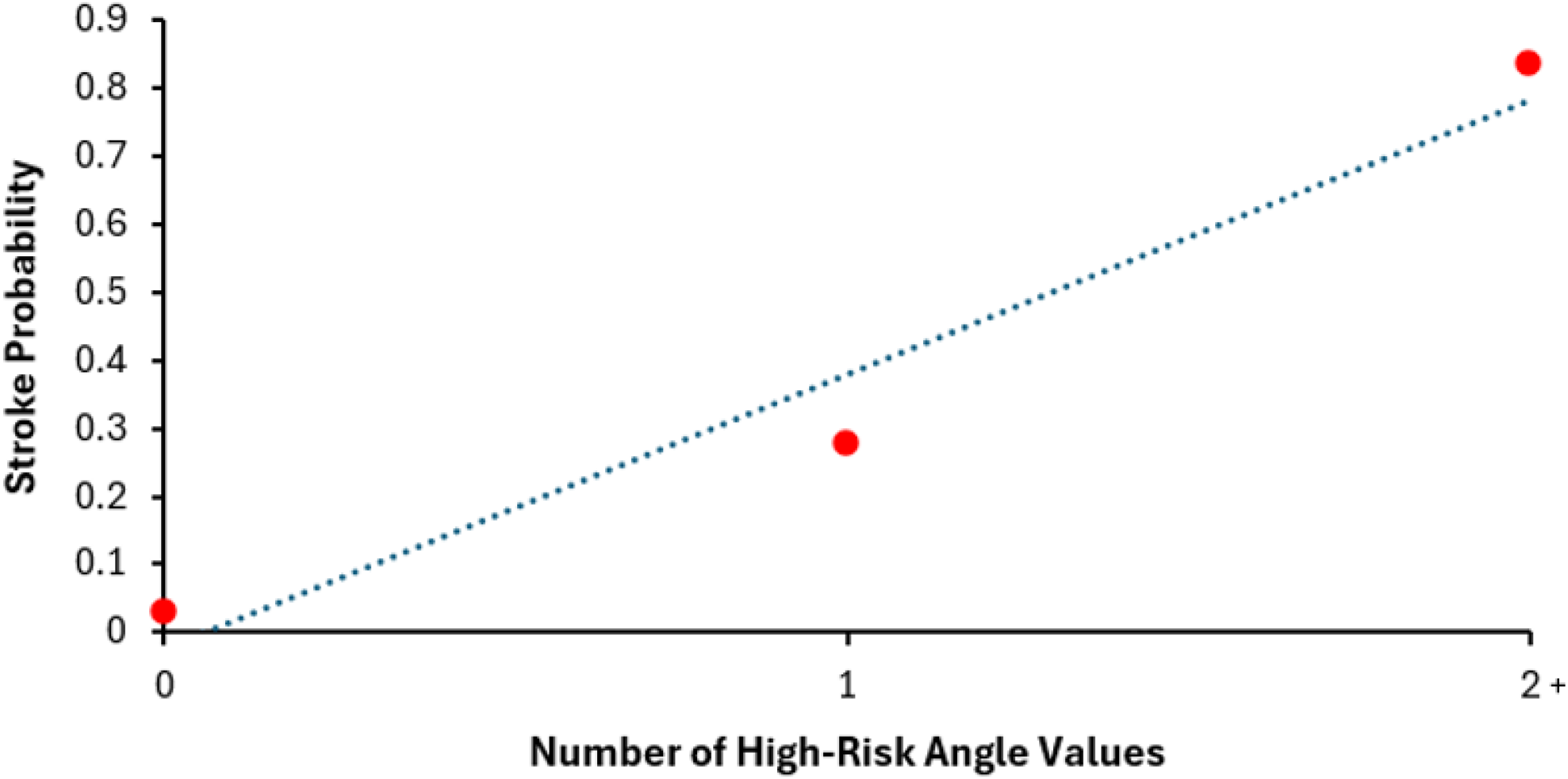
Stroke probability by the number of above-threshold angle values.

The predicted probability of stroke increased progressively with the number of high-risk angles, with patients in the 2+ group showing the highest predicted stroke probability compared to those with 0 or 1 high-risk angles. Specifically, patients without any high-risk angles had a stroke probability of only 2.8%. This increased to 27.7% for one high risk angle and then markedly increased to 83.4% for two or more high risk angles. The number of patients in each cohort and the concordance of their angle values with corresponding cutoffs are shown in Table 3. The positive and negative predictive values for each angioarchitectural parameter were calculated and are included in Table 4. No patients in the asymptomatic group had suffered a stroke at a mean follow-up of 47.2 months (range 24 - 77 months).

**Table 3.** Angle risk categorization for symptomatic and asymptomatic patients.

| Angle | Symptomatic Cohort (n=9) |  | Asymptomatic Cohort (n=26) |  |
| --- | --- | --- | --- | --- |
|  | n, high risk (%) | n, low risk (%) | n, high risk (%) | n, low risk (%) |
| IPT | 6 (66.7) | 3 (33.3) | 2 (7.7) | 24 (92.3) |
| CPT | 0 (0.0) | 9 (100.0) | 0 (0.0) | 26 (100.0) |
| IWP | 4 (44.4) | 5 (55.5) | 5 (19.2) | 21 (80.1) |
| CWP | 5 (55.5) | 4 (44.4) | 5 (19.2) | 21 (80.1) |

Abbreviations: CPT, common carotid artery – pouch-tip; CWP, common carotid artery – web-pouch; IPT, internal carotid artery – pouch-tip; IWP, internal carotid artery – web pouch

**Table 4.** Positive and Negative Predictive Values of Angioarchitectural Parameters.

| Angle | PPV (%) | NPV (%) |
| --- | --- | --- |
| IPT | 75.0 | 88.9 |
| CPT | 0.0 | 74.2 |
| IWP | 44.4 | 80.8 |
| CWP | 50.0 | 84.0 |

Abbreviations: CPT, common carotid artery – pouch-tip; CWP, common carotid artery – web-pouch; IPT, internal carotid artery – pouch-tip; IWP, internal carotid artery – web pouch; NPV, negative predictive value; PPV, positive predictive value

In the asymptomatic cohort, mean IWP (90.5°), CWP (40.8°), IPT (78.1°), and CPT (59.1°) angles all fell below the high-risk threshold values determined from our previous work. In the symptomatic cohort, mean IWP (96.9°), CWP (56.2°), and IPT (97.1°) angles were above the high-risk threshold values, whereas the mean CPT (53.6°) angle fell below the cut-off value of 89.4° for designation as a high-risk angle.

## Discussion

Our previous work used eleven symptomatic and eleven asymptomatic patients, to determine several parameters of CW angioarchitecture predictive of higher risk for experiencing future stroke. In this study, we validate this model within a significantly larger cohort of asymptomatic patients with incidentally discovered CWs and a fresh symptomatic cohort.

Our study found that within the group of asymptomatic patients with incidentally discovered CWs, the mean values for all four angles fell below the predetermined threshold values. Our results in this study, and in our previous work, have suggested that CPT and IWP angles are particularly useful for delineating high versus low stroke risk. Thus, these values may merit particular attention. However, while the mean angle values of the asymptomatic cohort fell below each cutoff value, outlier angle values above the threshold value were regularly seen. Similarly, symptomatic patients often have individual angle measurements that fall within the low stroke risk category. Thus, angioarchitectural parameters are more informative when considered as a group rather than when focusing on the value of a singular parameter.

For instance, if two or more of the four angle parameters for a given patient fall above their respective cutoff values, this is much more likely to indicate a significant risk of stroke than does having only a single high-risk value. In this manner, it may be more useful to consider how many angle values out of the four total fall above or below the threshold. Patients may be stratified into 0, 1, or 2+ high-risk angle categories, allowing for more distinct risk assignment. However, these values must also be interpreted within the unique clinical context of each patient, and the clinician must use their best judgment in management.

CWs remain an underrecognized etiology of ischemic stroke.^9^ This is in part due to their rarity, considering they are present in only 1.0-1.2% of all stroke patients.^2^ Additionally, in contrast to more common stroke etiologies like large atherosclerotic plaques, CWs are not traditionally associated with significant carotid stenosis and appear much more subtly on vascular imaging.^10^ Nonetheless, they have been demonstrated to significantly disrupt blood flow and can serve as a nidus for thrombus formation.^11^ For this reason, CWs have been said to increase the risk of stroke by a factor of 8.^12^ However, no previous studies have differentiated between symptomatic and asymptomatic CWs and their relative stroke risks.

One significant challenge in research investigating CW stroke risk is that the cause of CWs remains unknown. Thus, it is unclear if the difference between symptomatic and asymptomatic CWs is one solely of structure, as in the case of different geometric features (“angioarchitecture”), or if other factors, such as accumulated time with a CW, may also contribute to stroke. This is especially difficult considering it is undetermined whether CWs are present from birth or if they develop later in life.

Regardless, the differentiation between symptomatic and asymptomatic webs is crucial, as symptomaticity affects the next steps in clinical management and the determination of whether intervention is indicated. In the case of symptomatic webs, current evidence suggests that intervention is superior to conservative management, with one systematic review of 158 patients with CWs demonstrating a 56% recurrent stroke rate in patients receiving purely medical management versus a 0% recurrent stroke rate in those receiving surgical intervention.^13^

Carotid endarterectomy (CEA) is one intervention that may reduce the risk of stroke recurrence by resecting the carotid web, thereby clearing the ICA lumen.^14,15^ Therefore, continuing to develop and validate a model to grade stroke risk of incidentally discovered carotid webs may help identify high-risk patients who would benefit from intervention, such as CEA or carotid artery stenting (CAS), as opposed to conservative management. Similarly, patients with incidentally determined webs determined to have a low stroke risk may then avoid the morbidity associated with unnecessary surgery.

While this study’s results are encouraging and further validate this model of risk assessment, there are several limitations. First, all angle measurements are measured by hand, which makes this method prone to error from inter-user variability. While two investigators made all measurements in this study, there may still be some variability between them. A potential solution to this problem is the development of a machine learning image segmentation program that can measure these angles on CTA imaging in a consistent, standardized manner. This would also allow this method to be consistently applied by different providers and across different institutions.

This study is also limited by its small sample size; this model of risk assessment will require further validation and refinement within larger populations. Additionally, while our angioarchitectural parameters are meant to serve as proxies for flow dynamics around the CW, quantitative research is needed to determine if there are measurable flow dynamic differences between symptomatic and asymptomatic webs. Lastly, while the asymptomatic patients experienced no cerebrovascular events throughout their clinical follow-up, this period ranged from two to just over six years, and follow-up over a longer period may be necessary to definitively categorize patients as asymptomatic.

## Conclusions

Carotid webs with more high-risk angioarchitectural parameters on CTA, as defined by our previous work, can be used to identify a heightened risk for stroke. These angle values are most meaningful when assessed collectively for a patient. Having two or more high-risk angles is associated with a high probability of stroke. The carotid web stroke risk assessment strategy described in our previous work accurately categorized all asymptomatic patients with incidentally discovered carotid webs as low stroke risk across all four previously described parameters of carotid web angioarchitecture. Assessing geometrical carotid web parameters may guide risk stratification of patients with incidentally discovered webs, thereby improving the fidelity of surgical patient selection.

## Data Availability

The data are not publicly available to preserve patient confidentiality and due to institutional data sharing policies. However, the data are available from the corresponding author on reasonable request and with appropriate institutional approvals.???

## Acknowledgments

The authors wish to thank Karl L. Sangwon for the illustrations included in this article.

## Data Statement

The data are not publicly available to preserve patient confidentiality and due to institutional data sharing policies. However, the data are available from the corresponding author on reasonable request and with appropriate institutional approvals.

